# Rate discrimination training may partially restore temporal processing abilities from age-related deficits

**DOI:** 10.1101/2021.11.29.21266998

**Authors:** Samira Anderson, Lindsay DeVries, Edward Smith, Matthew J. Goupell, Sandra Gordon-Salant

**Affiliations:** Department of Hearing and Speech Sciences at University of Maryland, College Park, 20742, USA; Food and Drug Administration, 10903 New Hampshire Ave, Silver Spring, MD 20993-0002

**Author notes:** Corresponding Author: Samira Anderson, Ph.D. Department of Hearing and Speech Sciences 7251 Preinkert Dr. College Park, MD 20752.

**Keywords:** Auditory Training, Aging, Speech Perception, Auditory steady-state response

## Abstract

The ability to understand speech in complex environments depends on the brain’s ability to preserve the precise timing characteristics of the speech signal. Age-related declines in temporal processing may contribute to the older adult’s experience of communication difficulty in challenging listening conditions. This study’s purpose was to evaluate the effects of rate discrimination training on auditory temporal processing. A double-blind, randomized control design assigned 77 young normal-hearing, older normal-hearing, and older hearing-impaired listeners to one of two treatment groups: experimental (rate discrimination for 100-Hz and 300-Hz pulse trains) and active control (tone detection in noise). All listeners were evaluated during pre-and post-training sessions using perceptual rate discrimination of 100-, 200-, 300-, and 400-Hz band-limited pulse trains and auditory steady-state responses (ASSRs) to the same stimuli. Training generalization was evaluated using several temporal processing measures and sentence recognition tests that included time-compressed and reverberant speech stimuli. Results demonstrated a session × training group interaction for perceptual and ASSR testing to the trained frequencies (100 and 300 Hz), driven by greater improvements in the training group than in the active control group. Further, post-test rate discrimination of the older listeners reached levels that were equivalent to those of the younger listeners at pre-test. The training-specific gains generalized to untrained frequencies (200 and 400 Hz), but not to other temporal processing or sentence recognition measures. Further, non-auditory inhibition/attention performance predicted training-related improvement in rate discrimination. Overall, the results demonstrate the potential for auditory training to partially restore temporal processing in older listeners and highlight the role of cognitive function in these gains.

## Introduction

The brain’s ability to process the temporal characteristics of auditory stimuli is an integral component of speech understanding, particularly in complex environments that reduce the redundancy of the speech signal. For example, the ability to discriminate between changes in temporal rate contributes to the listener’s ability to discriminate fundamental frequency to cue speaker and gender identification, an important cue that supports speech segregation and speech understanding in noise. Previous studies have demonstrated age-related declines in rate discrimination (Gaskins et al. 2019) and in other temporal processing tasks (Fitzgibbons and Gordon-Salant 2011; Pichora-Fuller et al. 2007; Roque et al. 2019a). Therefore, temporal processing deficits may underlie older adults’ reported difficulties when communicating in challenging listening situations, and the question remains whether these age-related deficits can be improved through targeted auditory training.

Animal and human studies suggest that the brain retains some plasticity into older age; therefore, training that targets temporal tasks may improve perceptual performance and neural processing in the older listener. Age-related decreases in rat temporal coding and cortical firing synchrony can largely be reversed by training on a frequency discrimination auditory training paradigm (de Villers-Sidani et al. 2010). A cross-species study (mice and humans) found that adaptive training on signal-in-noise detection in a closed-loop paradigm led to improvements in signal detection in both species and generalization to speech-in-noise performance in human listeners (Whitton et al. 2017). Finally, a training study with older listeners, both with and without hearing loss, found that auditory-cognitive training led to reductions in latencies of the frequency-following response, an indication of improved temporal precision (Anderson et al. 2013). This training presented stimuli that adaptively increased or decreased both consonant-transition durations and auditory memory load. Overall, these studies demonstrate the potential for training-related neuroplasticity in older listeners.

The time course of perceptual learning and generalization to untrained stimuli has been compared across older and younger listeners (Manheim et al. 2018; Sabin et al. 2013). For example, Sabin et al. (2013) found differing learning patterns in older and younger listeners on a spectral modulation detection training task. Young listeners improved in their ability to detect spectral modulations, but this training effect did not generalize to an untrained spectral modulation frequency. In contrast, older listeners showed more modest and gradual improvement in performance throughout the training sessions that generalized to an untrained frequency. The authors surmised that a prolonged consolidation phase that stabilizes task learning may have facilitated this generalization.

Previous studies demonstrated improvement in rate discrimination thresholds in cochlear-implant listeners across a wide range of ages (Bissmeyer et al. 2020; Goldsworthy and Shannon 2014). However, it is currently unknown whether targeted auditory training can improve temporal rate discrimination ability in older acoustic normal-hearing listeners or hearing-impaired listeners, and whether improvement in temporal rate discrimination generalizes to performance on other temporal processing and speech understanding measures. The current study was designed to: 1) determine whether rate discrimination training can improve auditory temporal processing in older and younger listeners in both perceptual and in neural responses, 2) determine the extent to which perceptual learning on rate discrimination generalizes to other temporal processing tasks and measures of speech understanding, and 3) investigate the neural and cognitive variables that are associated with training-related improvements in perception. Based on previous animal and human studies, we hypothesized that perceptual training would partially restore temporal processing in older listeners. Furthermore, we hypothesized that neural responses to the trained pulse trains (auditory steady-state responses) and cognitive ability would relate to changes in perception. Finally, given that previous studies have not shown significant effects of hearing loss on temporal processing tasks (Fitzgibbons and Gordon-Salant 1996; Roque et al. 2019a), we hypothesized a similar training benefit regardless of hearing status.

## Materials and Methods

### Listeners

We recruited 301 listeners for a double-blind randomized controlled clinical trial and evaluated them to determine if they met the following audiometric criteria for these groups: young normal hearing (YNH, age 18-30 yrs), older normal hearing (ONH, age 65-85 yrs), and older hearing impaired (OHI, age 68-85 yrs). Normal hearing was defined as pure-tone thresholds ≤ 25 dB HL (re: ANSI, 2018) from 125 to 4000 Hz in the right ear and impaired hearing was defined by a high-frequency pure-tone average (average thresholds at 1, 2, and 4 kHz) > 30 dB HL and thresholds at 2 and 4 kHz < 70 dB HL (to ensure signal audibility). In all three listener groups, hearing thresholds were symmetrical (no interaural differences > 10 dB at any frequency), and there were no air-bone gaps > 10 dB at any frequency. Word recognition scores were > 70% bilaterally, using 25-word lists of the NU-6 test (Tillman and Carhart 1966) presented at 75 dB HL in quiet. Middle ear function was normal bilaterally based on age values for tympanometric peak pressure, peak admittance, tympanometric width, and equivalent volume; acoustic reflexes were present from 500-2000 Hz, elicited ipsilaterally and contralaterally. Finally, auditory brainstem responses (ABRs) were recorded, and Wave V latencies were < 6.8 ms with no interaural asymmetries > 0.2 ms. Additional criteria included the following: A passing score of ≥ 26 on the Montreal Cognitive Assessment (MoCA; Nasreddine et al. 2005), a negative history of neurological disease, a passing score on the Snellen vision screening chart ≤ 20/50 (Hetherington 1954), being a native English speaker, and earning a high school diploma. All procedures were reviewed and approved by the Institutional Review Board (IRB) at the University of Maryland, College Park. Participants provided informed consent and were monetarily compensated for their time.

The 125 listeners who met the study criteria were randomly assigned to one of two training groups: experimental and active control. Of these, 48 listeners did not complete the study. Seventeen listeners were dismissed due to: non-compliance with training (3), poor quality data (7), adverse event (1), and excessive time delay associated with COVID-19 (6). Twenty-six listeners withdrew from the study due to medical or transportation issues. Eleven listeners were lost to follow-up. The final numbers of listeners in each training group were 40 Experimental (14 YNH, 16 ONH, and 10 OHI; 30 Females) and 37 Active Control (15 YNH, 14 ONH, and 8 OHI; 28 Females). See Table 1 for additional demographic characteristics. Note that across measurements, 1% of listener data (31 of 2618 measurements) are missing because of isolated issues during data collection or because of anomalous data that did not converge.

**Table 1.** Demographic characteristics of Experimental and Active Control groups including Sex, Age, and High-Frequency Pure-Tone Average (HF PTA). YNH = young normal hearing, ONH = older normal hearing, OHI = older hearing impaired, F = female, M = mean, and S.D. = standard deviation.

| Group | Experimental |  |  | Active Control |  |  |
| --- | --- | --- | --- | --- | --- | --- |
|  | YNH<br>(n=14) | ONH<br>(n=16) | OHI<br>(n=10) | YNH<br>(n=15) | ONH<br>(n=14) | OHI<br>(n=8) |
| Sex | 7 F | 15 F | 8 F | 9 F | 12 F | 7 F |
| Age | 21.1 (M) | 69.9 (M) | 74.0 (M) | 21.0 (M) | 70.0 (M) | 74.4 (M) |
|  | 2.2 (S.D) | 4.0 (S.D) | 6.4 (S.D) | 2.0 (S.D) | 4.5 (S.D) | 6.4 (S.D) |
| HF PTA | 5.5 (M) | 14.1 (M) | 38.2 (M) | 6.2 (M) | 13.9 (M) | 36.8 (M) |
|  | 2.6 (S.D) | 3.0 (S.D.) | 4.4 (S.D.) | 2.9 (S.D) | 4.3 (S.D.) | 7.1 (S.D.) |

### Pre- and Post-Testing

Both training groups were tested using the same battery of electrophysiologic and behavioral measures prior to the onset and after completion of training. Auditory steady-state responses (ASSRs) were recorded to 100-, 200-, 300- and 400-Hz bandpass-filtered click trains, and behavioral pulse-rate discrimination was measured to the same stimuli. The behavioral test battery also included generalization measures: gap detection, gap duration discrimination, tempo discrimination, and several speech recognition measures. These measures will be described in more detail below.

#### Procedure

Listeners were seated in a double-walled sound-attenuating booth. The stimuli were presented to listeners through a single insert earphone (ER-2, Etymotic, Elk Grove Village, IL). Stimulus presentation and event timing were controlled from a laptop computer and a custom MATLAB script.

### Perceptual and Neural Responses to Pulse Trains

#### Stimuli

The stimuli were band-limited pulse trains (300-ms duration) having rates of 100, 200, 300, and 400 Hz. The pulses had a 1-kHz bandwidth arithmetically centered around 4 kHz, created using forward-backward Butterworth filters (5^th^ order) (Gaskins et al. 2019). Raised cosine Hanning windows with a 10-ms rise-fall time were applied to the stimuli to avoid filter-related onset and offset transients. The stimuli were presented monaurally to the right ear at 75 dBA for all neural and non-speech perceptual measures described below. For perceptual testing only, a low-frequency masking noise was mixed with the pulse train stimuli to eliminate the use of low-frequency distortion products to perform the task. Wideband masking noise was low-pass filtered using a 200-Hz cutoff with a −3 dB/octave filter and presented at an overall level of 61 dB SPL.

### Perceptual Rate Discrimination

Rate discrimination for each reference pulse rate was assessed by measuring pulse-rate difference limens (DLs) using a three-interval, two-alternative forced choice (3I-2AFC) procedure. Each rate (100, 200, 300, and 400 Hz) was tested with three blocks of 60 trials for a total of 720 trials across blocks. The order of reference pulse rate was randomized.

The listeners viewed a monitor that displayed four boxes. Stimulus presentation was self-paced throughout the experiment. They were asked to click the box containing “Begin Trial” and then heard a sequence of three stimuli, with the presentation of each stimulus synchronized to a flash in the corresponding visual block in the sequence. The first stimulus was always the reference stimulus. The target stimulus with the higher rate was in the second or third interval, randomly chosen with a 50% a priori probability.

The listeners received the following instructions: “You will hear three brief sounds that sound like a buzz. The first one is the ‘standard.’ One of the other sounds has a slightly higher pitch that sounds different from the standard sound. Please select the sound, 2 or 3, that contains the higher pitch (or sounds different from the standard sound). If you are not sure, take a guess.”

After each listener response, correct answer feedback was provided by flashing a green light at the box corresponding to the correct interval. A 2-down-1-up adaptive procedure was employed to target 70.7% correct on the psychometric function (Levitt 1971). The initial rate difference between the reference and target stimulus was set at 40%. The maximum allowable rate difference was 40% and the minimum allowable rate difference was 0% (i.e., adaptive tracks could not go below the reference rate). The adaptation step size was then decreased by a factor of 2 until the listener reached three reversals, after which the step size decreased by a factor of √2.

#### Analysis

Perceptual responses were recorded in MATLAB. The pulse rate difference limen (DL) in percent for an individual adaptive track was found by calculating the geometric mean over all of the reversals in the adaptive procedure except the first two. The arithmetic mean of the second and third tracks was used to calculate the final DL for each listener and condition. The first track was omitted to decrease the effects of learning from the first track. The DLs were log-transformed due to a negative skew in the data prior to conducting the statistical analysis.

### ASSR

#### Recording

The pulse trains were presented at a rate of 1.66 Hz using the Intelligent Hearing Systems Continuous Acquisition Model (IHS SEPCAM, Miami, FL) through electromagnetically shielded insert ER-3 earphones (IHS) in an electrically shielded double-walled sound-attenuating booth. A three-electrode vertical montage was used (Cz active, right ear lobe reference, low forehead ground). Responses were recorded with a 10-kHz sampling rate and were filtered from 1 to 5 kHz on-line. A minimum of 1024 artifact-free sweeps (≤ 30 µV) were obtained for each condition. The listeners watched their movie of choice, muted with subtitles, to facilitate a relaxed but awake state.

#### Data Analysis

Responses were imported into MATLAB format using the pop_biosig function from EEGLAB (Delorme and Makeig 2004) and filtered from 50-500 Hz. An individual average response was created with the first 1000 artifact-free sweeps. Phase-locking factor (PLF) was assessed in a manner similar to that employed in previous studies (Jenkins et al. 2018; Roque et al. 2019b), using Morlet wavelets to decompose the signal from 50-500 Hz (Tallon-Baudry et al. 1996). The PLF value was then calculated for the response time region of 10-310 ms and around a 20-Hz frequency bin corresponding to the pulse-rate of each condition. The PLFs were log-transformed due to a negative skew in the data.

### Gap Detection

Gap detection thresholds were measured using target stimuli that were 250-ms wideband Gaussian noise bursts that had a silent gap temporally centered in the stimulus. Cosine squared windows with a 1-ms rise-fall time were applied to the stimuli to avoid transients.

A 3I-2AFC procedure was used. The first interval was the standard, with no gap. The target stimulus with the silent gap was in the second or third interval, randomly chosen with a 50% *a priori* probability.

The listeners received the following instructions, “This is the ‘standard’ and is a continuous noise. One of the other noise bursts, 2 or 3, has a very brief pause or interruption that sounds different from the standard noise burst. Please select the noise burst, 2 or 3, that contains the brief pause (or sounds different from the standard tone pair). If you are not sure, take a guess.”

After each listener response, correct answer feedback was provided. Then the gap duration was adapted according to the 2-down-1-up adaptive rule, targeting 70.7% correct discrimination. The initial gap duration was 25 ms. The maximum gap duration was 100 ms and the minimum gap duration was 1 ms. The initial step size in the adaptive procedure was 5 ms. After two reversals, the step size was changed to 1 ms. The adaptive track continued until there were eight reversals. Threshold was defined as the arithmetic mean of the last six reversals. Three adaptive tracks were conducted. The arithmetic mean of the second and third tracks was used to calculate the gap detection threshold for each listener.

### Gap Duration Discrimination

Gap duration discrimination was measured using 250-ms 1000-Hz tone pairs separated by a silent interval (Fitzgibbons and Gordon-Salant 1994). Cosine squared windows with a 5-ms rise-fall time were applied to the stimuli to avoid transients.

The listener received the following instruction: “Please select the tone pair, 2 or 3, that contains the longer silent interval (or sounds different from the standard tone pair). If you are not sure, take a guess.”

After each listener response, correct answer feedback was provided. Then the gap duration was adapted according to the 2-down-1-up adaptive rule. The initial gap duration for the target was 350 ms (i.e., 40% larger than the reference gap of 250 ms). The maximum gap duration was 450 ms and the minimum gap duration was 252 ms. The initial step size in the adaptive procedure was 10 ms. After two reversals, the step size was reduced to 2 ms. The adaptive track continued until there were eight reversals. The relative duration discrimination DL in percent (based on the 250-ms reference) was calculated from the arithmetic mean of the last six reversals. Three adaptive tracks were measured. The arithmetic mean of the second and third tracks was used to calculate the gap duration discrimination DL for each listener.

### Tempo (Rhythm) Discrimination

Discrimination DLs were measured for inter-onset intervals (IOIs) in isochronous sequences of five brief 50-ms 1000-ms tones (see Fitzgibbons and Gordon-Salant 2001). The IOI is defined as the duration between the onset of one tone in the sequence and the onset of the subsequent tone. Cosine squared windows with a 5-ms rise-fall time were applied to the stimuli to avoid transients.

A 3I-2AFC procedure was used. The reference intervals had a fixed IOI, either 100 ms (fast reference) or 600 ms (slow reference). The target stimulus with the relatively slower tone sequence was in the second or third interval, randomly chosen with a 50% *a priori* probability.

The listeners received the following instructions: “You will hear three sequences of 5 brief tones. The first sequence is the ‘standard.’ One of the other sequences, 2 or 3, sounds slower than the standard sequence. Please select the tone sequence, 2 or 3, that is a slower sequence (or sounds different from the standard sequence). If you are not sure, take a guess.”

After each listener response, correct answer feedback was provided. Then the IOI was adapted according to the 2-down-1-up adaptive rule. The starting target IOI was 150 ms for the 100-ms reference IOI and 700 ms for the 600-ms reference IOI. The maximum target IOI was 200 and the minimum target IOI was 101 ms for the 100-ms reference IOI; the maximum target IOI was 800 and the minimum target IOI was 601 ms for the 600-ms reference IOI. The initial step size in the adaptive procedure was 10 ms. After two reversals, the step size decreased to 2 ms. The adaptive track continued until there were eight reversals. The DL for each IOI was calculated from the arithmetic mean of the last six reversals of each track. Three adaptive tracks were conducted for each reference IOI (i.e., there were six separate adaptive tracks). The arithmetic mean of the second and third tracks was used to calculate the relative IOI DL in percent (based on either the 100-ms or 600-ms IOI reference) for each listener.

### Sentence Recognition

Sentence recognition in quiet was measured for sentences from the IEEE corpus (IEEE 1969) in five conditions: normal rate with no reverberation, two levels of time compression (40% and 60%) and two levels of reverberation (0.6 s and 1.2 s). There were 10 sentences presented in each condition. Each sentence was preceded by a carrier phrase, “Number 1,” “Number 2,” etc. Listeners were instructed to repeat the sentence they heard. The experimenter scored which of the five key words in each sentence were repeated correctly, and the percent correct keywords words out of 50 was calculated for each condition.

### Training

#### Experimental

Listeners received in-lab perceptual rate-discrimination training for two rates, 100 and 300 Hz, using a procedure similar to that described above for rate discrimination assessment. The training was blocked by rate, with four blocks of 60 trials for each rate for a total of 480 trials. Correct-answer feedback was provided after each trial throughout the training sessions. Nine sessions of this training took place in the sound-attenuating booth over the course of two to three weeks.

#### Active Control

Listeners received in-lab training on tone-in-noise detection, using a 3I-2AFC procedure. A notched-noise paradigm and simultaneous masking were used to measure filter bandwidths (Desloge et al., 2012), using a 300-ms 1-kHz stimulus tone and a 500-ms white Gaussian noise (0.25-6 kHz). The target tone was temporally centered in the noise. Cosine squared windows with a 10-ms rise-fall time were applied to the noise and target tones to avoid transients. The noise level was fixed at 75 dBA and the tone level varied adaptively to determine threshold in three notch bandwidths: 90, 120, and 150 Hz.

After each listener response, correct answer feedback was provided. Then the tone level was adapted according to the 2-down-1-up adaptive rule. The initial target tone level was 75 dBA. The maximum target tone level was 75 dBA and the minimum target tone level was −20 dBA. The initial step size in the adaptive procedure was 3 dB. After three reversals, the step size decreased to 0.5 dB. Each of the three notch bandwidth conditions was presented in four blocks, with 40 trials per block, for a total of 480 trials; therefore, the procedure had the same number of trials when compared to the pulse-rate discrimination training, except that the task was different. Nine sessions of this training took place in the sound-attenuating booth over the course of two to three weeks. The masked threshold in dB for an individual adaptive track was found by calculating the arithmetic mean over the last four reversals in the adaptive track. The arithmetic mean of the second and third tracks was used to calculate the final masked threshold for each listener and condition.

### Cognitive Testing

Assessments from the National Institutes of Health (NIH) Cognition Toolbox (Weintraub et al. 2013) were used to determine if particular cognitive skills predicted perceptual training benefits. These tests included the List Sorting Working Memory Test, the Flanker Inhibitory Control and Attention Test, the Pattern Comparison Processing Speed Test, and the Dimensional Card Sort Test. The tests were administered using the NIH toolbox application on an Apple iPad (Apple, Inc., Cupertino, CA). The uncorrected standardized scores were downloaded from the application.

### Statistical Analysis

#### Pulse Rate Discrimination Improvement and Near Generalization

A four-way repeated measures analyses of variance (RMANOVA) was conducted to evaluate the effects of training on perception and neural representation of the 100- and 300-Hz pulse trains, comparing pre-test and post-test measures. There were two between group-variables (listener group and training group) and two within-group variables (rate: 100 and 300 Hz; session: pre-test vs. post-test). The dependent variable was pulse-rate DL for perceptual testing. A separate four-way RMANOVA was performed with the dependent variable PLF for the ASSR.

To assess near generalization to untrained rates (200 and 400 Hz), two separate four-way RMANOVAs were conducted for perceptual testing and ASSR using the same variables. To account for differences in learning stemming from pre-test performance (Sabin et al. 2013), we conducted a one-way repeated-measures analysis of covariance (RMANCOVA) using post-test 100- and 300-Hz DLs as the dependent variables and the pre-test 100- and 300-Hz DLs as covariates. In addition, multivariate ANOVAs were conducted to assess differences between post-testing rate discrimination in the older listeners with pre-testing rate discrimination in the YNH listeners to determine if training restores temporal processing deficits in the ONH listeners. Bonferroni-corrected independent-samples t tests and paired-samples t tests (assuming equal variance) were used to perform post hoc analyses when main effects or interactions were observed.

#### Mid Generalization

Separate three-way RMANOVAs were conducted to evaluate mid generalization to the other temporal processing measures as follows, using the same between-group variables (listener group and training group) as for the pulse trains and the same within-subject variable of session (pre-test and post-test). Dependent variables for two of the measurements were gap detection threshold and gap duration discrimination DL. For the tempo discrimination experiment, the dependent variable was IOI discrimination DL. In addition, there was an additional within-group variable (reference IOI: 100 and 600 ms), making this a four-way RMANOVA.

#### Far Generalization

A RMANOVA was conducted to evaluate generalization to sentence recognition measures using the same between-group variables. The one within-group variable was condition (Clean speech, two levels of time compression, and two levels of reverberation), and the other was test time (pre-test, post-test). The dependent variable was the sentence recognition score. The percent scores were transformed using the rationalized arcsine (rau) transform proposed by Studebaker (1985) to avoid violation of the homogeneity of variance assumption required for an ANOVA.

#### Performance Predictors

A step-wise multiple linear regression was conducted to identify the potential factors that contributed to changes in pulse-rate discrimination performance for 100- and 300-Hz rates in the experimental group. The dependent variable was the average change in rate DL (post - pre) for 100- and 300-Hz reference rates. Processing speed was included as an independent variable due to its relationship to pre-test DLs (Gaskins et al. 2019). Additional cognitive measures included in the analyses were working memory, the Dimensional Card Sort (cognitive flexibility), and the Flanker (attention and inhibitory control). The PTA in the right ear (500 to 4000 Hz) was also included to determine the contribution of audibility to performance. A log transform was used to normalize the skewed PTA distribution. Finally, to determine the contributions of subcortical neural processing to performance changes, the pre-test PLF and change in PLF averaged for 100- and 300-Hz rates were included.

## Results

### Trained rates (100 and 300 Hz)

Figure 1 displays pre- and post-test performances for the 100- and 300-Hz reference rates in YNH, ONH, and OHI listeners. The RMANOVA showed a main effect of session (*F*_(1,69)_ = 59.33, *P* < 0.001, *η^2^* = 0.42), such that DLs were lower (better) at the post-test compared to the pre-test. There was a significant training group × session interaction (*F*_(1,69)_ = 5.48, *P* = 0.022, *η^2^* = 0.04), There was a main effect of session in both the experimental group (*F*_(1,37)_ = 39.20, *P* < 0.001, *η^2^* = 0.51) and the active control group (*F*_(1,35)_ = 22.63, *P* < 0.001 *η^2^* = 0.35), but a larger effect size and more pronounced DL decreases were noted in the experimental group (100 Hz: 56%; 300 Hz: 86%) than in the active control group (100 Hz: 19%; 300 Hz: 56%). Therefore, although there was a procedural learning effect in both groups, the interaction between the training groups suggests additional perceptual learning in the experimental group that exceeded the procedural learning effect. The training group × listener group × session interaction was not significant (*F*_(2,69)_ = 1.38, *P* = 0.258, *η^2^* = 0.02), suggesting that training effects on rate discrimination did not differ significantly by listener group. A RMANCOVA using post-test 100- and 300-Hz DLs as the dependent variables and the pre-test 100- and 300-Hz DLs as covariates confirmed greater effects of training (lower post-training DLs) in the experimental group than in the active control group (*F*_(1,71)_ = 9.59, *P* = 0.003, *η^2^* = 0.08).

**Figure 1.**
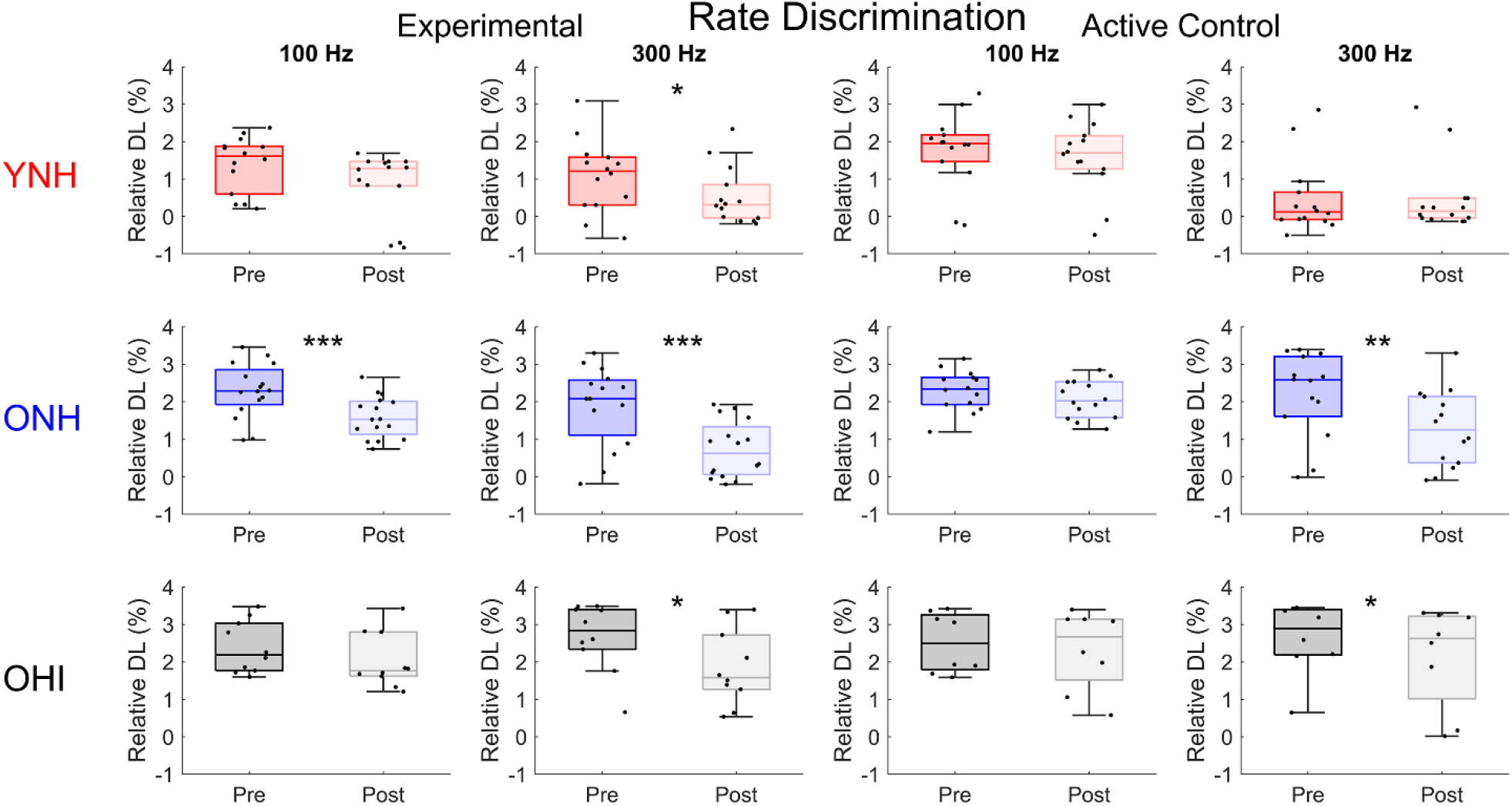
Rate discrimination at two training rates. Box plots and individual data points displaying pre- and post-training relative difference limens (DL) as a function of 100- and 300-Hz rates obtained in young normal-hearing (YNH), older normal-hearing (ONH), and older hearing-impaired (OHI) listeners who completed nine sessions of rate-discrimination training (experimental group) or tone-in-noise detection training (active control group). Note that these percentages are log-transformed. There were significant improvements in performance (smaller DLs) in the experimental group that were not observed in the active control group. *P <0.05, **P <0.01, ***P <0.001. Medians: Inside box lines. Upper and lower quartiles: top and bottom edges of the box, respectively. The endpoints of the whiskers represent the range of values without the outliers.

There was also a significant effect of rate (*F*_(1,69)_ = 35.74, *p* < 0.001, *η^2^* = 0.26), because there were lower DLs for the 300-compared to the 100-Hz rate. There was a main effect of listener group (*F*_(2,69)_ = 23.53, *P* < 0.001, *η^2^* = 0.39), such that the YNH listeners had lower DLs than the ONH (*P* < 0.001) and OHI (*p* < 0.001) listeners, but the ONH and OHI listeners did not significantly differ (*P* = 0.087). In addition, there was a significant listener group × rate interaction (*F*_(2,69)_ = 17.03, *P* < 0.001, *η^2^* = 0.24). There was a significant listener group difference for the 300-Hz rate (*F*_(2,71)_ = 7.12, *P* = 0.002, *η^2^* = 0.17) but not for the 100-Hz rate (*F*_(2,72)_ = 0.91, *P* = 0.408, *η^2^* = 0.03).

### Untrained rates (200 and 400 Hz)

Figure 2 displays pre-test and post-test performance for the 200- and 400-Hz pulse rates in YNH, ONH, and OHI listeners. The RMANOVA showed a main effect of session (*F*_(1,70)_ = 21.24, *P* < 0.001, *η^2^* = 0.19), such that DLs were lower at the post-test compared to the pre-test. There was a significant training group × session interaction (*F*_(1,70)_ = 8.01, *P* = 0.006, *η^2^* = 0.07). There was a significant main effect of session in the experimental group (*F*_(1,37)_ = 21,48, *P* < 0.001, *η^2^* = 0.33) that was not present in the active control group (*F*_(1,35)_ = 2.59, *P* = 0.117, *η^2^* = 0.06). The training group × listener group × session interaction was not significant (*F*_(2,70)_ = 0.28, *P* = 0.754, *η^2^* = 0.005), suggesting that training effects on rate discrimination did not differ significantly by listener group. A RMANCOVA using post-test 200- and 400-Hz DLs as the dependent variables and the pre-test 200- and 400-Hz DLs as covariates confirmed greater effects of training (lower post-training DLs) in the experimental group than in the active control group (*F*_(1,68)_ = 13.53, *P* < 0.001, *η^2^* = 0.10).

**Figure 2.**
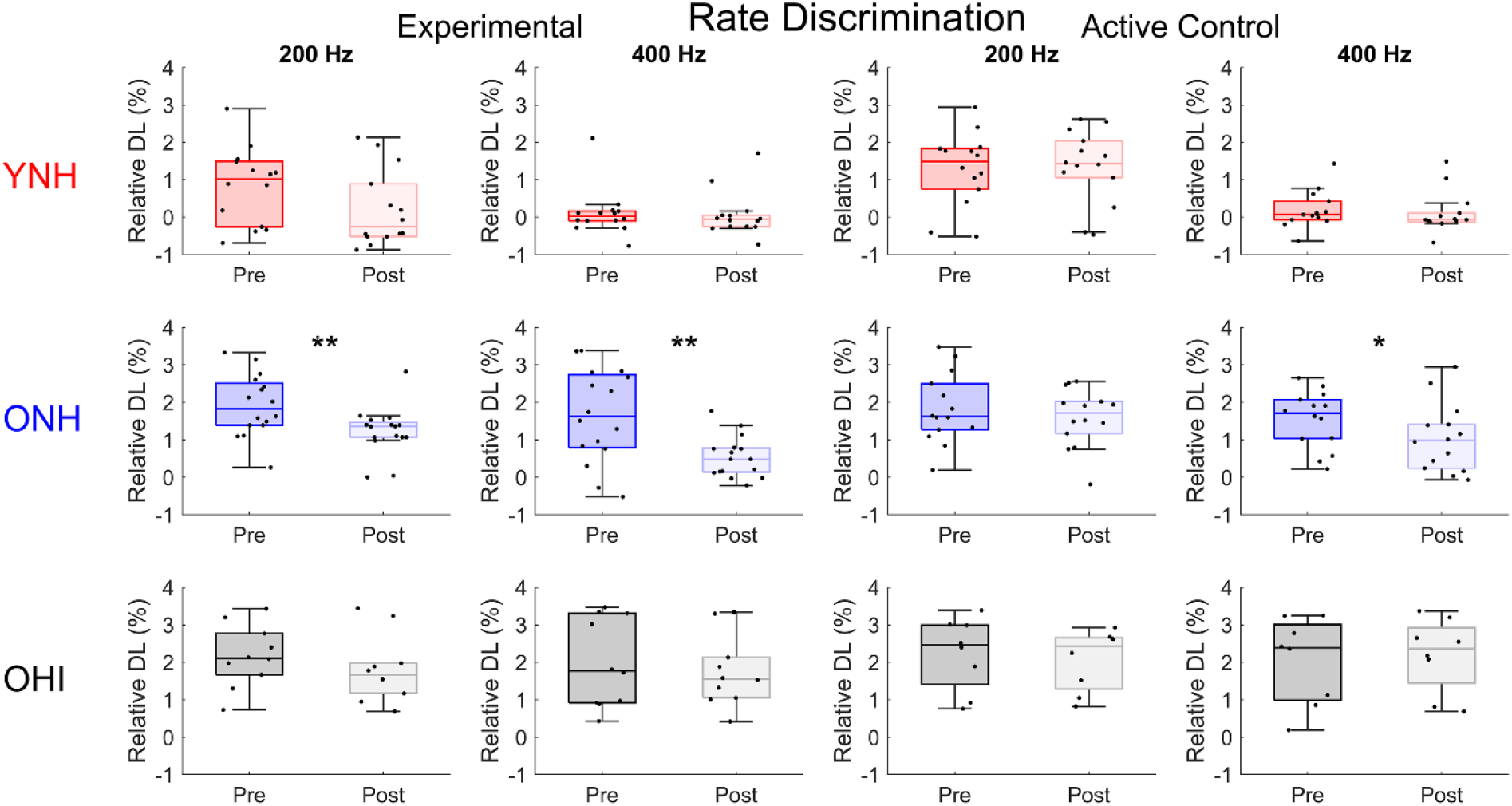
Rate discrimination at two untrained rates. Box plots and individual data points displaying difference limens (DLs) as a function of 200- and 400-Hz rates obtained in young normal-hearing (YNH), older normal-hearing (ONH), and older hearing-impaired (OHI) listeners who completed nine sessions of rate discrimination training (experimental) or signal detection in noise training (active control). Note that these percentages are log-transformed. There were significant improvements in performance (decreased DLs) in the experimental group (especially the ONH and OHI), that were not observed in the active control group (except for the ONH 400-Hz rate). *P <0.05, **P <0.01. Medians: Inside box lines. Upper and lower quartiles: top and bottom edges of the box, respectively. The endpoints of the whiskers represent the range of values without the outliers.

The RMANOVA showed a significant effect of rate (*F*_(1,69)_ = 29.45, *P* < 0.001, *η^2^* = 0.25) associated with lower DLs for the 400-Hz rate than the 200-Hz rate. There was a main effect of listener group (*F*_(2,69)_ = 28.70, *P* < 0.001, *η^2^* = 0.41), such that the YNH listeners had lower DLs than the ONH (*P* < 0.001) and OHI (*P* < 0.001) listeners, and the ONH listeners had lower DLs than the OHI listeners (*P* = 0.039). The rate × listener group interaction was also significant (*F*_(2,70)_ = 6.45, *P* = 0.003, *η^2^* = 0.11), driven by larger listener group differences for the 400-Hz rate compared to the 200-Hz rate.

A multivariate analysis of variance (MANOVA) was then used to compare the post-test DLs in the ONH and OHI listeners to the pre-test DLs in the YNH listeners in the experimental training group for the four different rates (Figure 3). At the pre-test, there was a main effect of listener group (*F*_(2,36)_ = 14.28, *P* < 0.001, *η^2^* = 0.44); both groups of older listeners had higher (poorer) DLs than the YNH listeners (*P* < 0.001), but the older groups did not differ from each other (*P* > 0.99). A comparison of the pre-test YNH DLs with the post-test DLs in ONH and OHI listeners showed a main effect of listener group (*F*_(2,37)_ = 8.29, *P* = 0.001, *η^2^* = 0.31), but post hoc t tests showed that the DLs of ONH listeners did not differ from those of YNH listeners (*p =* 0.426), while the OHI listeners had higher DLs than both the ONH (*p =* 0.025) and the YNH (*p <* 0.001) listeners. There was also a rate × listener group interaction (*F*_(6,111)_ = 4.68, *P* < 0.001, *η^2^* = 0.13). At the 100-Hz rate, there were no significant differences among the three listener groups (*P* = 0.18. At the 200-, 300-, and 400-Hz rates, there was no significant difference between the YNH and ONH listeners (*P* > 0.05), but the OHI listeners had higher DLs than the YNH listeners (*P* < 0.05). Given that pre-test DL differences existed between the ONH and YNH listeners (*P <* 0.001), these results demonstrate that training on rate discrimination at least partially restored temporal processing abilities on this measure in ONH listeners.

**Figure 3.**
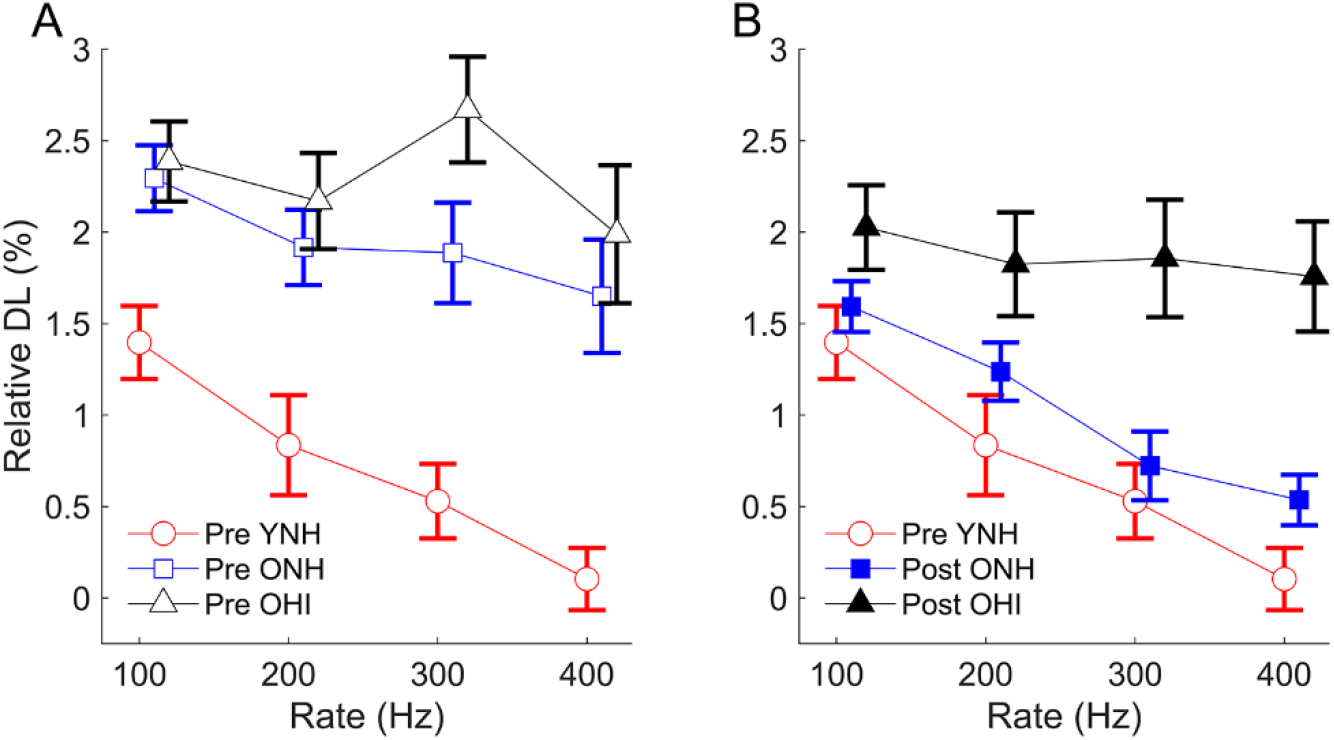
Relative DLs (log-transformed) are compared between young normal-hearing (YNH), older normal-hearing (ONH), and older hearing-impaired (OHI) experimental training groups for pre-test data (Panel A) and between pre-test YNH and post-test ONH and OHI groups (Panel B). The pre-test differences between YNH and ONH groups were not present at post-test, but differences persisted for the OHI groups. Errors bars: ± 1 S.E.

### ASSR

#### Trained rates (100 and 300 Hz)

Figure 4 displays pre- and post-training box plots and average PLFs for the 100- and 300-Hz rates measured from the YNH, ONH, and OHI listeners. The RMANOVA showed a training group × session interaction (*F*_(1,69)_ = 6.63, *P* = 0.012, *η^2^* = 0.08), driven by a significant increase in PLF in the experimental group (*F*_(1,35)_ = 5.14, *P* = 0.03, *η^2^* = 0.11) that was not observed in the active control group (*F*_(1,34)_ = 2.01, *P* = 0.165, *η^2^* = 0.05). The training group × listener group × session interaction was not significant (*F*_(1,69)_ = 0.09, *P* = 0.916, *η^2^* = 0.002), suggesting that training effects on PLF did not differ by listener group. To account for differences in neuroplasticity effects stemming from pre-test performance, we conducted a RMANCOVA using post-test 100- and 300-Hz PLFs as the dependent variables and the pre-test 100- and 300-Hz PLFs as covariates and confirmed greater effects of training (higher post-training PLFs) in the experimental group than in the active control group (*F*_(1,68)_ = 8.89, *P* = 0.004, *η^2^* = 0.079).

**Figure 4.**
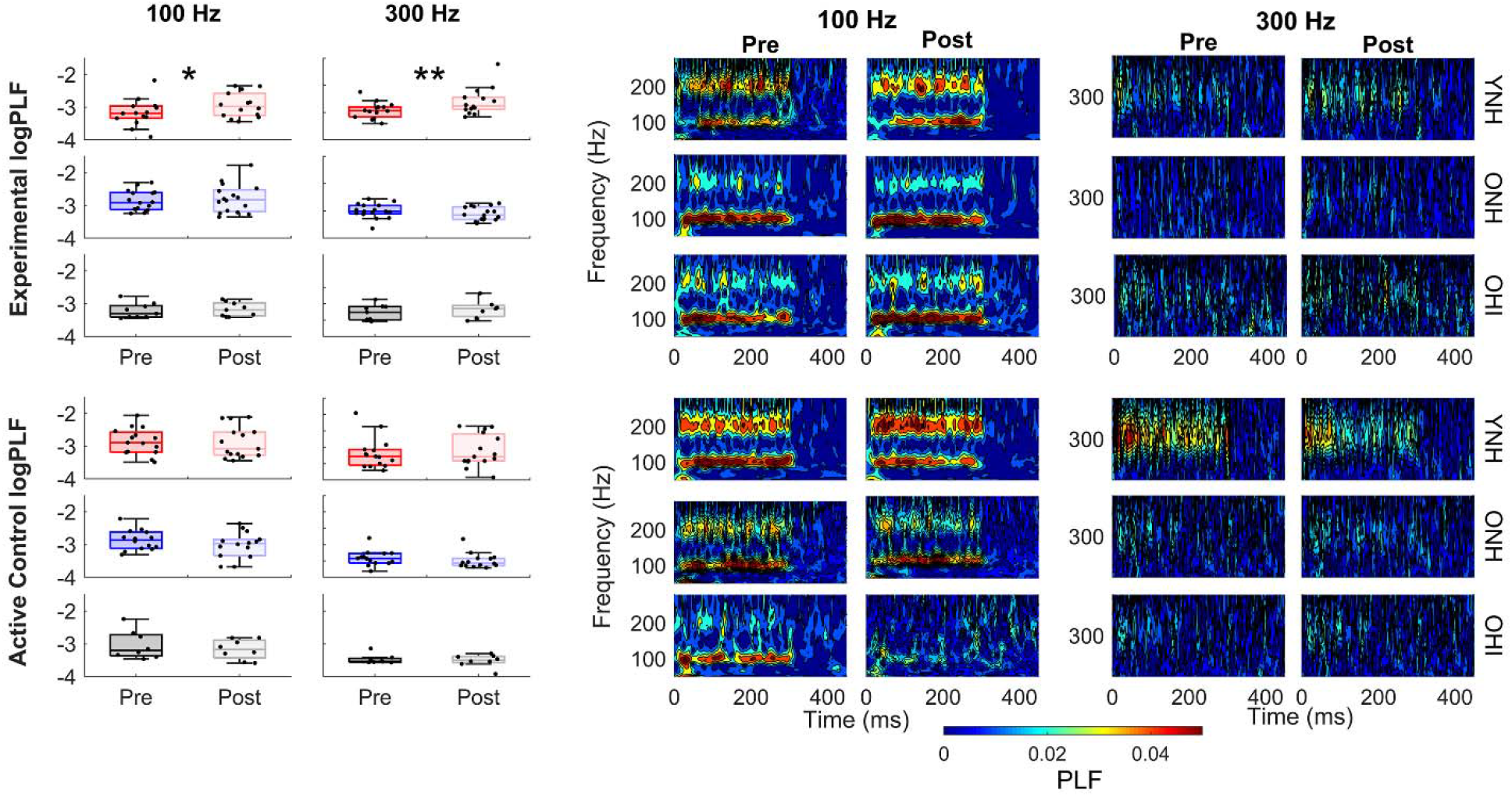
Pre- and post-training phase-locking factor (PLF) for 100- and 300-Hz rates is displayed in box plots and in the time-frequency domain for young normal-hearing (YNH), older normal-hearing (ONH), and older hearing-impaired (OHI) listeners in the experimental (top three panels) and active control (bottom three panels) groups. Note that these values are log-transformed. There were significant increases in PLF in the training group, especially in the YNH listeners, that were not observed in the active control group. *P <0.05, **P <0.01. Medians: Inside box lines. Upper and lower quartiles: top and bottom edges of the box, respectively. The endpoints of the whiskers represent the range of values without the outliers.

The RMANOVA showed a significant effect of rate (*F*_(1,69)_ = 82.49, *P* < 0.001, *η^2^* = 0.51) associated with higher PLFs for the 100-Hz rate than the 300-Hz rate. There was no main effect of listener group (*F*_(2,69)_ = 1.67, *P* = 0.195, *η^2^* = 0.04), but there was a significant listener group × rate interaction (*F*_(2,69)_ = 4.10, *P* = 0.021, *η^2^* = 0.05). There was no significant listener group difference for the 100-Hz PLF (*F*_(2,73)_ = 0.85, *P* = 0.431, *η^2^* = 0.02), but there was a significant group difference for the 300-Hz PLF (*F*_(2,72)_ = 6.86, *P* = 0.002, *η^2^* = 0.16). Post hoc t tests showed that the YNH group had higher PLFs than the ONH group (*P* = 0.002), but the group difference was not significant between the YNH and OHI groups (*P* = 0.057) nor between the ONH and OHI groups (*P* = 0.866).

#### Untrained rates (200 and 400 Hz)

Figure 5 displays pre- and post-training box plots and average PLFs for the 200- and 400-Hz rates for the YNH, ONH, and OHI listeners. The RMANOVA showed that the training group × session interaction was not significant (*F*_(1,68)_ = 0.99, *P* = 0.356, *η^2^* = 0.01) and there was no main effect of session (*F*_(1,68)_ = 0.002, *P* = 0.96, *η^2^* < 0.001), suggesting that training effects on PLF did not generalize to untrained rates.

**Figure 5.**
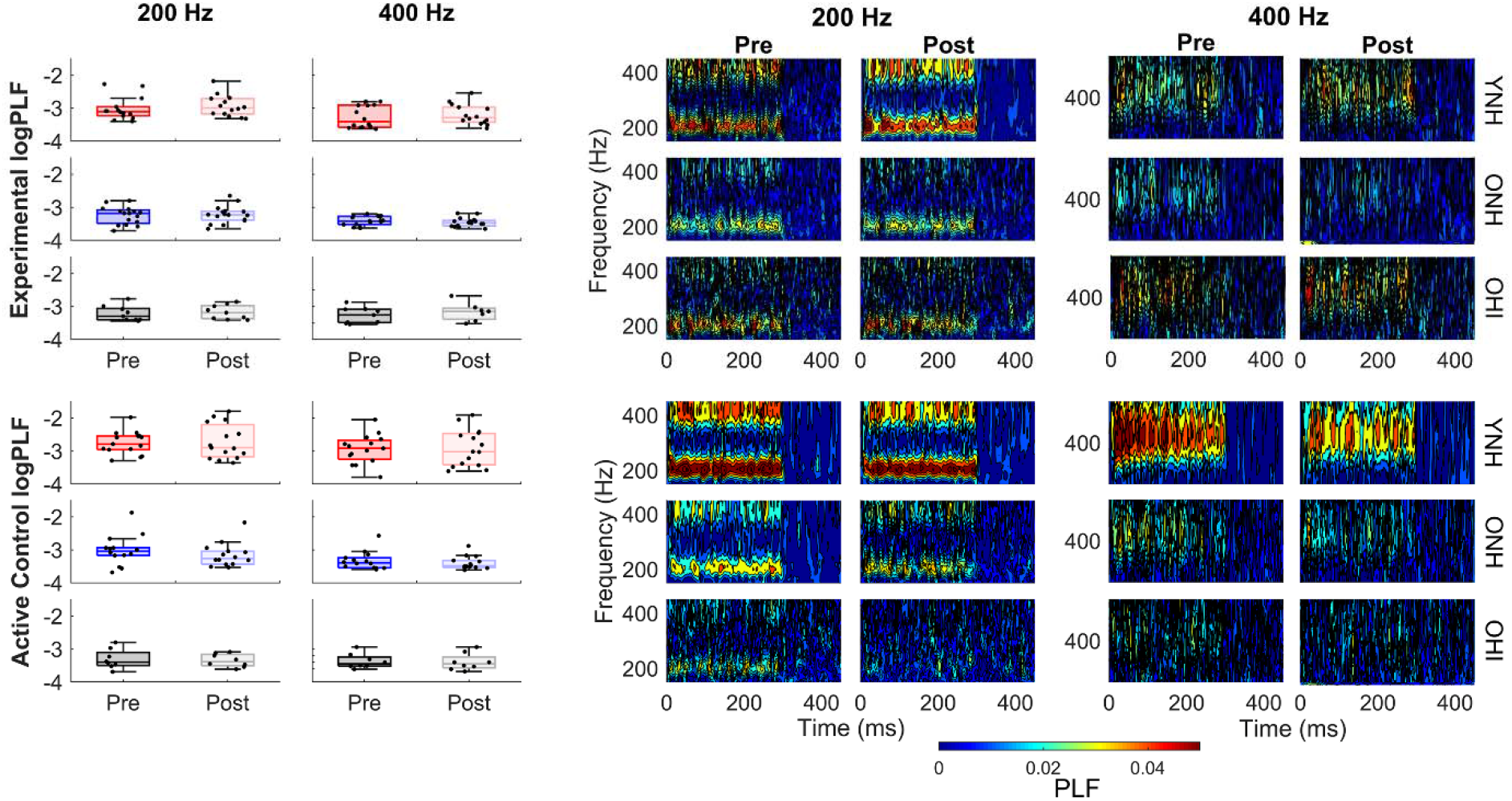
Pre- and post-training phase-locking factor (PLF) for 200- and 400-Hz rates is displayed in box plots and in the time-frequency domain for young normal-hearing (YNH), older normal-hearing (ONH), and older hearing-impaired (OHI) listeners in the experimental (top three panels) and active control (bottom three panels) groups. No increases in PLF were noted in any group. Medians: Inside box lines. Upper and lower quartiles: top and bottom edges of the box, respectively. The endpoints of the whiskers represent the range of values without the outliers.

There was a significant effect of rate (*F*_(1,68)_ = 25.30, *P* < 0.001, *η^2^* = 0.25) associated with higher PLF for the 200-Hz rate than the 400-Hz rate. There was a main effect of listener group (*F*_(2,68)_ = 15.16, *P* < 0.001, *η^2^* = 0.28), such that the YNH listeners had higher PLFs than either the ONH (*p* < 0.001) or OHI (*P* < 0.001) listeners, but there were no significant differences between the ONH and OHI listeners (*P* = 1.00). In addition, there was a significant listener group × rate interaction (*F*_(2,68)_ = 3.34, *P* = 0.04, *η^2^* = 0.07), driven by larger listener group differences for the 200-Hz than for the 400-Hz rate.

### Mid Generalization – Temporal Processing

#### Gap Detection and Gap Duration Discrimination

Figure 6 displays pre- and post-training box plots and individual datapoints for the gap detection and gap duration discrimination tasks in YNH, ONH, and OHI listeners.

**Figure 6.**
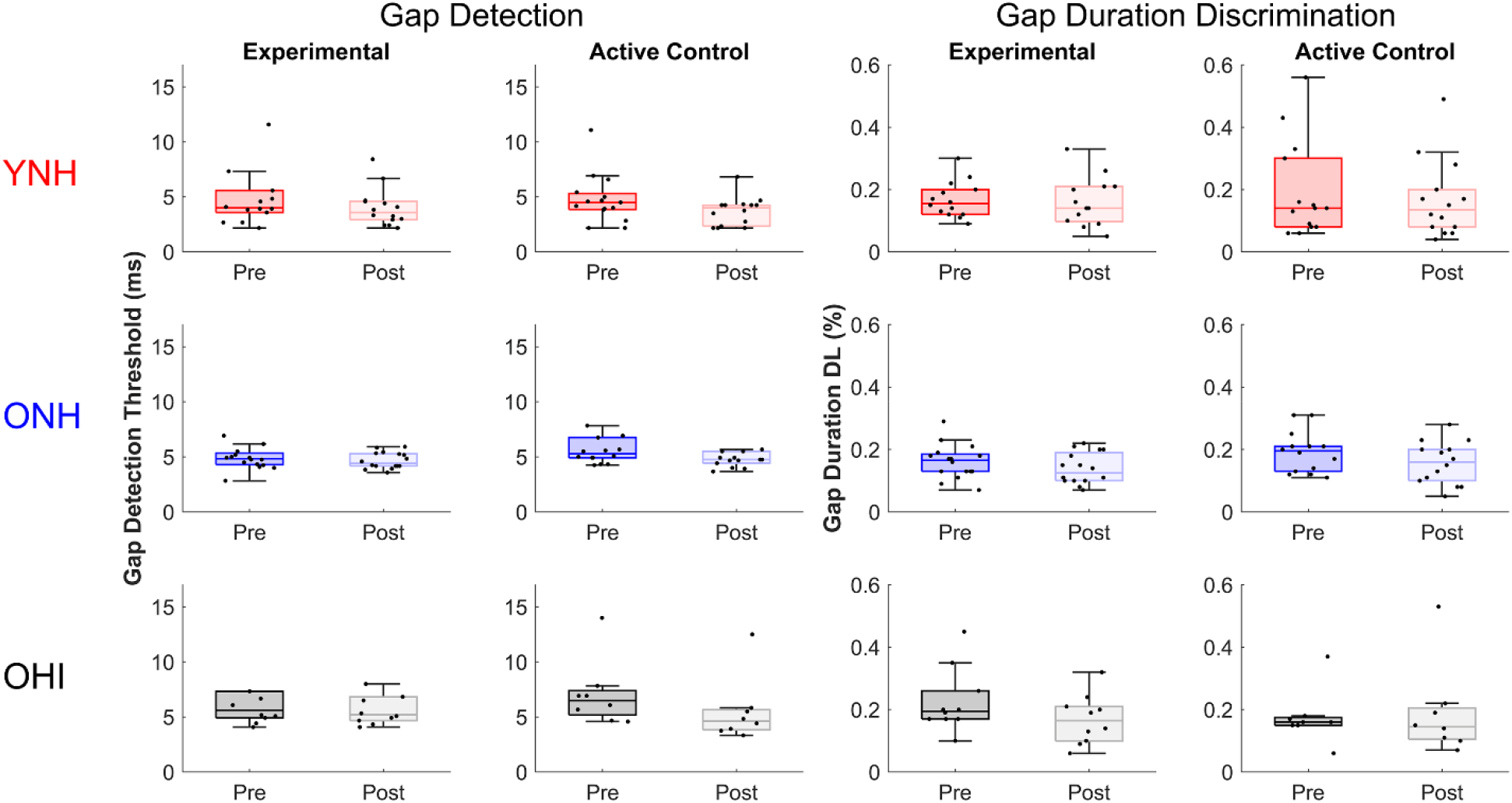
Pre- and post-training gap detection thresholds and gap duration DLs are displayed in box plots and individual data points for young normal-hearing (YNH), older normal-hearing (ONH), and older hearing-impaired (OHI) listeners in the experimental and active control groups. No changes in performance were noted from pre-test to post-test in any group. Medians: Inside box lines. Upper and lower quartiles: top and bottom edges of the box, respectively. The endpoints of the whiskers represent the range of values without the outliers.

##### Gap detection

The RMANOVA showed that there was a main effect of session (*F*_(1,70)_ = 5.41, *P* = 0.023, *η^2^* = 0.01), but there was no training group × session interaction (*F*_(1,70)_ = 0.09, *P* = 0.77, *η^2^* < 0.01). There was no main effect of listener group (*F*_(2,70)_ = 1.51, *P* = 0.29, *η^2^* = 0.03).

##### Gap duration discrimination

The RMANOVA showed a main effect of session (*F*_(1,69)_ =7.00, *P* = 0.01, *η^2^* = 0.01), but there was no training group × session interaction (*F*_(1,69)_ = 0.56, *P* = 0.46, *η^2^* < 0.01). The was no main effect of listener group (*F*_(2,69)_ = 0.29, *P* = 0.75, *η^2^* < 0.01).

### Tempo Discrimination

Figure 7 displays pre- and post-training box plots and individual datapoints for relative DLs as a function of 100- and 600-ms IOIs in YNH, ONH, and OHI listeners.

**Figure 7.**
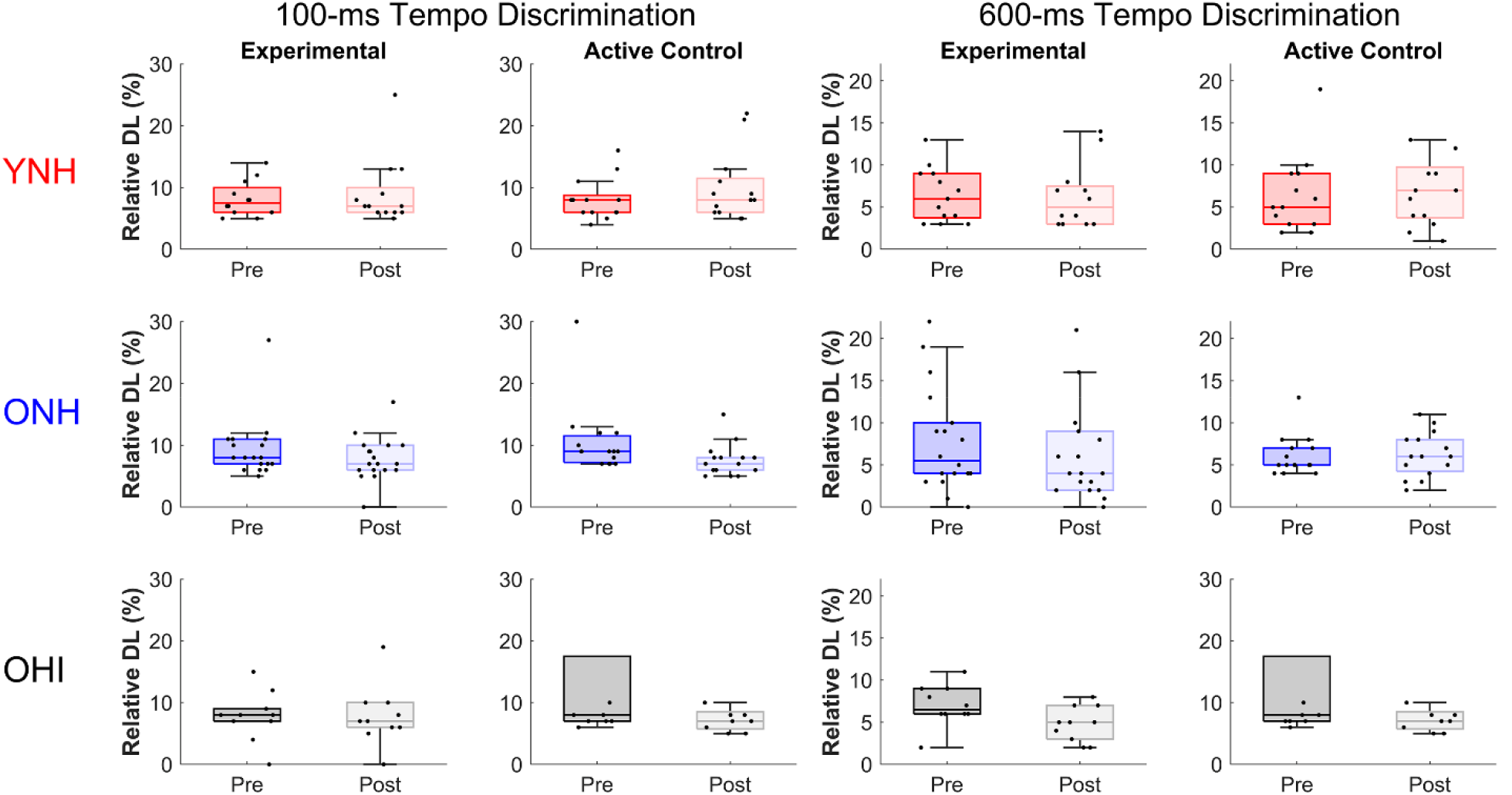
Box plots and individual data points for relative difference limens (DL) as a function of 100- and 600-ms inter-onset intervals (IOIs) obtained in young normal-hearing (YNH), older normal-hearing (ONH), and older hearing-impaired (OHI) listeners in the experimental and active control groups. No changes in performance were noted in any group. Medians: Inside box lines. Upper and lower quartiles: top and bottom edges of the box, respectively. The endpoints of the whiskers represent the range of values without the outliers.

The RMANOVA showed that there was no main effect of session (*F*_(1,66)_ = 1.10, *P* = 0.301, *η^2^* = 0.02) nor a training group × session interaction (*F*_(1,66)_ = 0.02, *P* = 0.893, *η^2^* < 0.001). The was no main effect of listener group (*F*_(2,66)_ = 0.36, *P* = 0.696, *η^2^* = 0.01). There was a main effect of IOI (*F*_(1,66)_ = 23.66, *P* < 0.001, *η^2^* = 0.23); the relative DLs were smaller for the 600-ms IOI than for the 100-ms IOI. No other interactions were significant.

### Speech Recognition

Figures 8 and 9 display pre- and post-training speech recognition data in experimental and active control groups, respectively. The RMANOVA showed that there was no main effect of session (*F*_(1,72)_ = 1.10, *P* = 0.299, *η^2^* < 0.01), nor a training group × session interaction (*F*_(1,71)_ = 0.77, *P* = 0.381, *η^2^* < 0.01), suggesting that sentence recognition did not improve across groups. There was a main effect of listener group (*F*_(2,71)_ = 60.03, *P* < 0.001, *η^2^* = 0.29). Post hoc testing showed that the OHI listeners had poorer overall performance than the YNH and ONH listeners (*P* < 0.001 for both), and ONH listeners had poorer overall performance than the YNH listeners (*P* = 0.008). There was a significant measure × listener group interaction (*F*_(8,284)_ = 44.82, *P* < 0.001, *η^2^* = 0.08). This interaction was driven by greater effects of time compression and reverberation on the performance of the OHI listeners than on that of the YNH or ONH listeners. Removal of the outlier in the OHI experimental group did not change these results.

**Figure 8.**
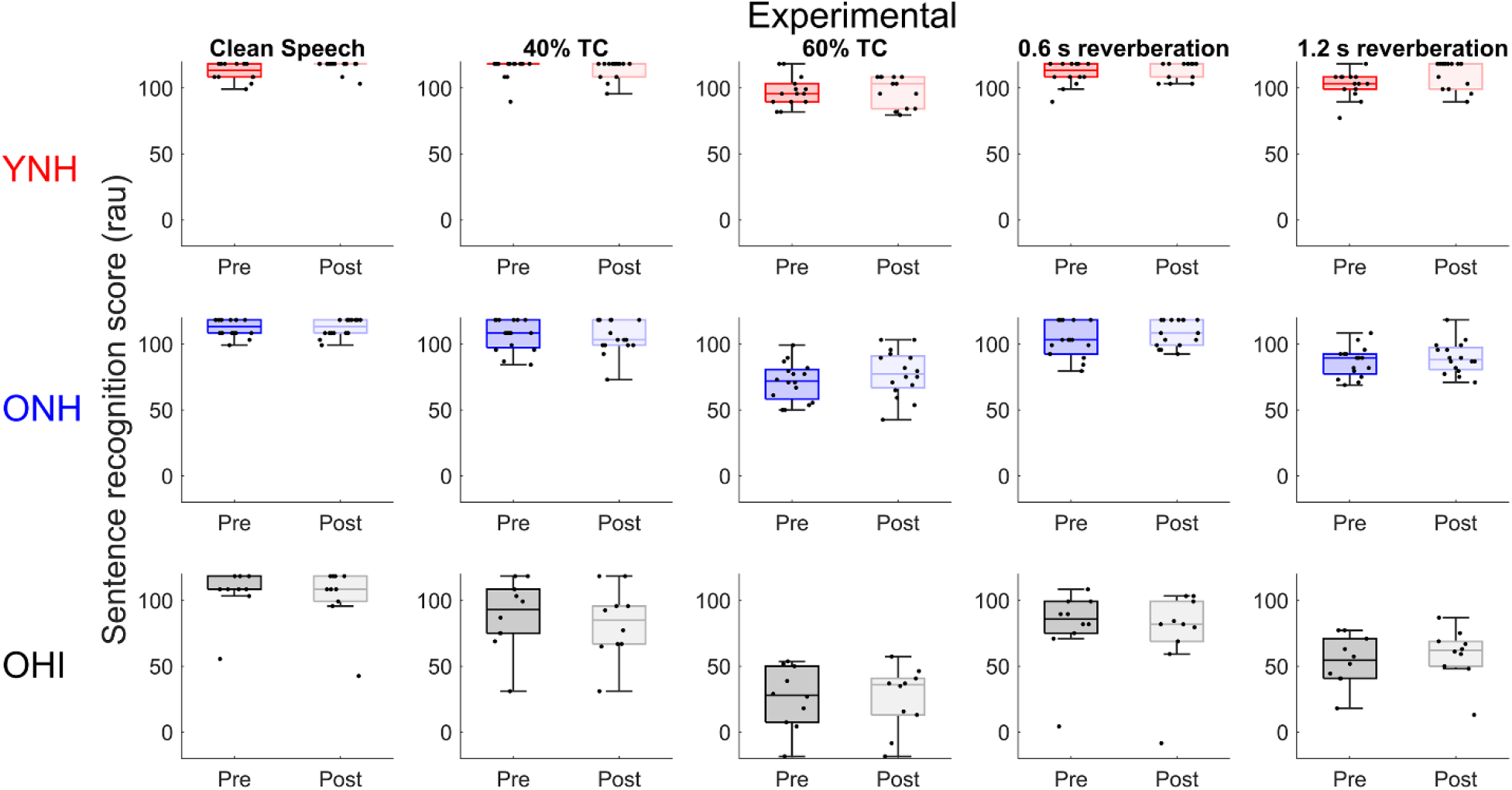
Experimental group. Box plots and individual data points displayed for rau-transformed percent of correct items pre- and post-training for clean (undistorted) speech, 40% time-compressed speech (40% TC), 60% time-compressed speech (60% TC), and 0.6 s and 1.2 s reverberation time in young normal-hearing (YNH), older normal-hearing (ONH), and older hearing-impaired (OHI) listeners in the experimental group. No changes in performance were noted in any listener group. Medians: Inside box lines. Upper and lower quartiles: top and bottom edges of the box, respectively. The endpoints of the whiskers represent the range of values without the outliers. Rau: rationalized arcsine transform

**Figure 9.**
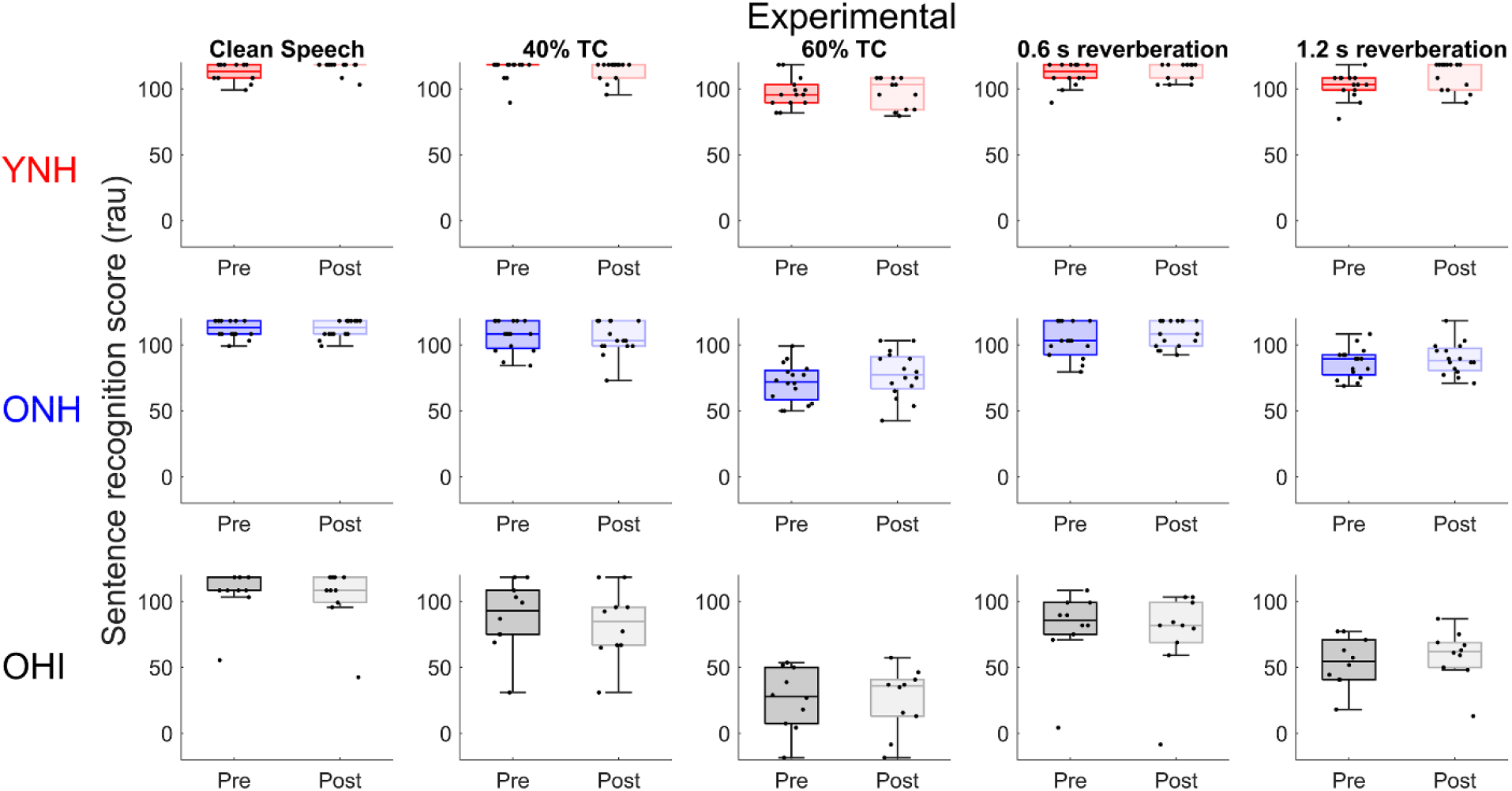
Active control. Box plots and individual data points displayed for rau-transformed percent of correct items pre-and post-training for clean speech, 40% time-compressed speech (40% TC), 60% time-compressed speech (60% TC), and 0.6 s and 1.2 s reverberation time in young normal-hearing (YNH), older normal-hearing (ONH), and older hearing-impaired (OHI) listeners in the active control group. No changes in performance were noted in any listener group. Medians: Inside box lines. Upper and lower quartiles: top and bottom edges of the box, respectively. The endpoints of the whiskers represent the range of values without the outliers. Rau: rationalized arcsine transform

### Factors Contributing to Training-Induced Changes in Pulse Rate Discrimination

The multiple linear regression collinearity diagnostics showed satisfactory tolerance (lowest 0.30) and variance inflation factor (highest 2.61) values, suggesting that the predictor variables were not highly correlated. One significant regression equation was returned; the Flanker score (attention) significantly predicted change in rate discrimination (*F*_(1,35)_ = 13.53, *P* < 0.001) with an *R^2^* value of 0.29. None of the other variables contributed significantly to the change in rate discrimination. This model is summarized in Table 2.

**Table 2.** Unstandardized (B) and standard error (S.E.) coefficients and standardized (β) coefficients in a model automatically generated by evaluating the significance of each variable’s contribution to the average change in 100- and 300-Hz rate discrimination. Only one model was generated, in which the Flanker score predicts significant variance in rate discrimination change. All other variables were excluded from the model (Working memory, speed of processing, dimension card sort, pure-tone average, pre-training phase-locking factor, and change in phase-locking factor).

| Variable | R<sup>2</sup> change | B | S.E. | β | 95% C.I. for B | p value |
| --- | --- | --- | --- | --- | --- | --- |
| Model 1 | 0.35 |  |  |  |  | <0.001 |
| Flanker |  | 0.26 | 0.06 | 0.59 | 0.14-0.39 | <0.001 |

## Discussion

The overarching goal of this investigation was to determine the effect of rate discrimination training on temporal processing in older and younger listeners. The results showed training-related improvements in temporal rate discrimination DLs and phase locking, but improvement did not generalize to other temporal processing tasks or sentence recognition measures. A smaller degree of improvement in temporal rate discrimination DLs was also noted in the active control group, likely an effect of procedural learning (Koziol and Budding 2012). The training × listening group interactions were not significant, suggesting that training effects were not specific to a specific listener group.

### Effects of Training in Older and Younger Listeners

Although the magnitude of change in DL appeared to be greatest in the ONH listeners, there was no significant listener × training group interaction, suggesting that training effects did not differ by age or hearing loss status (Fig. 1). These results appear to contrast with those of Sabin et al. (2013), who found differences in perceptual learning patterns between YNH and OHI listeners. The older listeners in the Sabin et al. study had mild to moderate hearing loss (thresholds ranging from 15 to 70 dB HL from 0.5 to 4 kHz), which may have affected their ability to benefit from training on spectrotemporal modulation due to decreased spectral resolution associated with hearing loss. Our study focused on a measure of temporal processing, an acoustic dimension that is less affected by hearing loss (Fitzgibbons and Gordon-Salant 1996), and we did not find effects of hearing loss on pre-training rate discrimination.

One important finding of our study was the improvement in behavioral temporal processing with training to partially reduce age-related deficits. The ONH listeners’ post-training DLs decreased to levels that were approaching those of the YNH listeners’ pre-training DLs (Fig. 3). These results are consistent with animal models of neuroplasticity in auditory aging that have shown that perceptual training can reduce or eliminate age-related deficits in temporal processing (de Villers-Sidani et al. 2010). However, we did not find a similar reduction of the age-related deficit in neural temporal processing. Significant group differences in the PLF (at rates > 100 Hz) at the pre-test session persisted at the post-test session. Our selection of rates was motivated to match testing between rate discrimination and the ASSR, and rates of 100-400 Hz arise from low to high brainstem sources (Herdman et al. 2002). The de Villers-Sidani study found changes in temporal precision in the rat auditory cortex, and therefore it is possible that a selection of a lower frequency rate (40 Hz or lower) that represents cortical sources would have shown an improvement in temporal precision.

### Generalization

#### Rate discrimination

Generalization of training effects was limited to “near generalization;” in other words, to discrimination of the untrained 200- and 400-Hz rates (Fig. 2). Although the session × listener × training group interaction was not significant, the listener × training group interaction was significant for ONH listeners (*p* = 0.04), but not for the YNH or OHI listeners (*p* > 0.05), suggesting that generalization was specific to the ONH listeners. The lack of generalization in the YNH listeners is consistent with previous studies that have found limited generalization effects for training on spectromodulation detection (Sabin et al. 2012) and amplitude-modulation detection (Fitzgerald and Wright 2011). Also, in YNH listeners, Wright et al. (2010) found that performance improved after two days of training (900 trials per day) on a temporal-interval discrimination task for a 1000-Hz tone pip and a 100-ms interval. Our training entailed 240 trials on each of the trained rates per day, and even though training occurred over 9 days, perhaps a greater number of trials per day is required to instill generalization in the YNH listeners. The spectromodulation detection task employed by Sabin et al. (2012) and Sabin et al. (2013) employed 720 trials during each training session. In older listeners, improvement in spectromodulation detection on the 2-cycles per octave trained condition generalized to the 1-cycle per octave untrained condition (Sabin et al. 2013). The authors observed that the most learning in YNH listeners occurred early in the training, but the ONH listeners exhibited a more prolonged time course of learning that may facilitate generalization. A similar phenomenon may underlie the generalization observed in ONH listeners in the current study.

#### ASSR

No generalization was found for untrained rates (200 and 400 Hz). The absence of generalization suggests two points: 1) the lack of increased PLF to 200- and 400-Hz rates suggests that the increase to 100- and 300-Hz rates is due to effects of training rather than to the effects of repeated testing, and 2) cortical neural processes may underlie generalization in perceptual performance, but the ASSR recordings in the current study targeted subcortical processing.

#### Generalization to other temporal processing and sentence recognition measures

No far generalization was observed for any of the temporal processing or sentence recognition measures. This is in contrast to other training studies employing temporally based training that have observed generalization to speech stimuli. For example, Lakshminarayanan and Tallal (2007) trained YNH listeners’ perception of frequency-modulated (FM) sweeps that varied in direction of change, duration of FM sweep, and inter-stimulus interval between sweeps. They found that this training led to enhanced discrimination between syllables that differed in the onset of the second formant (/ba/ vs /da/), transition duration (/ba/ vs/ /wa/), and silence duration (/sa/ vs /sta/). The transfer of temporally based training has also been observed in older listeners. Fostick et al. (2020) trained older listeners with normal to mild hearing loss levels on a spatial temporal order judgement task and found that improvement on this task generalized to recognition of word stimuli presented in quiet, narrowband noise, and wideband noise. They did not observe similar generalization for training on an intensity discrimination task. They interpreted these results as supporting the hypothesis that increased temporal processing ability leads to improvement in speech recognition.

Other training studies employing speech stimuli have observed generalization, and these effects vary depending on training parameters (Banai and Lavner 2019; Burk and Humes 2008; Karawani et al. 2015). Banai and Lavner (2019) trained young listeners to recognize time-compressed sentences under several different listening protocols that varied by stimulus set size, training schedule (trials presented in one training session vs. several sessions), and training duration. They found that all protocols led to improvement on the trained task and generalization to untrained tasks (new talker or sentences), but training over several sessions was the only protocol that led to generalization to new untrained sentences. The authors concluded that distributed training provides multiple opportunities to consolidate learning. Therefore, the use of speech rather than non-speech stimuli (as in the current study) may provide more opportunities for consolidation of learning due to the possibility of encountering similar stimuli in the natural environment.

### Factors that Contribute to Perceptual Learning

The Flanker score was the only variable that contributed significantly to change in rate discrimination from pre-test to post-test. Individuals with better response inhibition/attention experience greater decreases in relative DLs following training. We had initially hypothesized that both cognitive and ASSR measures would relate to changes in rate discrimination. This hypothesis was based in part on the results of Gaskins et al. (2019), who found that both processing speed and ASSR spectral energy predicted 400-Hz rate discrimination. The current study found relationships among all of the cognitive variables and the pre-test relative DLs (*r^2^* values ranging from 0.14 to 0.37), but not among the pre-test ASSR PLFs and relative DLs (no *r^2^* value higher than 0.10). Overall, the current results suggest that cognitive function could be an important factor in the potential for improvement in temporal processing ability, at least with respect to rate discrimination. We note that the relatively high rates used in the current study arise from brainstem sources (Herdman et al. 2002). Perhaps the inclusion of a lower rate emanating from the cortex (e.g., ≤ 40 Hz) would reveal a relationship between ASSR PLF and perceptual change due to the likelihood that cortical sources may be more highly influenced by top-down cognitive influences.

### Conclusion

The current results suggest that perceptual training improves rate discrimination across listeners and can partially restore behavioral auditory temporal processing deficits in older listeners. Neural phase locking also improves with training, but there was no relationship among behavioral and neural measurements with the tested rates. At least one measure of cognitive function, response inhibition/attention, accounts for significant variance in improvement in rate discrimination. Therefore, the paradigm used in the study protocol may be efficacious for individuals with average attention ability, but individuals with impaired attention or cognitive function may benefit from a different paradigm.

## Data Availability

All data produced in the present study are available upon reasonable request to the authors

## Acknowledgments

We would like to acknowledge the P01 Project 2 team (Graduate assistants: Alyson Schapira, Rachel Zimmerman, Abigail Poe, Logan Fraser, Mary Zhou, Jennifer Borja, Sydney Hancock, Alexandra Papanicolau, Andrew Morris, and Amarachukwa Ezenwa; Human Subject Research Coordinator: Carol Gorham) for dedication and hard work, and we thank Beverly Wright for her suggestions regarding the training and testing protocols. We would also like to thank the NIH National Institute on Aging for funding this project (P01 5P01AG055365).

## Notes

### Competing Interest Statement

The authors have declared no competing interest.

### Clinical Trial

NCT03475043

### Funding Statement

This study was funded by the National Institute on Aging (P01AG055365 to Gordon-Salant)

### Author Declarations

The Institutional Review Board of the University of Maryland gave ethical approval for this work.

